# Padel Sites in Metropolitan France: Spatial and Sociodemographic Characterization of Residents’ Potential Noise Exposure

**DOI:** 10.1101/2025.10.08.25337575

**Authors:** Jean-Charles Dufour, Christophe Bonnet

## Abstract

**Background:** The rapid expansion of padel in France has raised concerns about its coexistence with nearby residents due to its distinctive acoustic signature, characterized by impulsive, repetitive sounds. Guidance often proposes minimum setbacks from dwellings, yet no national-scale quantification of at-risk configurations has been available. We map where proximity and visibility combine to raise potential exposure and estimate affected populations.

**Methods:** Using aggregated open sources (878 sites), two independent examiners classified sites into three potential exposure classes based on distance and the presence of built screens (κ=0.83). GIS analyses then characterised, for at-risk (Class 1) sites, residential buildings within 100 m, their line-of-sight (LoS) to a reference court, and distance bands of 0–50 m and 50–100 m. Populations and households were estimated via dasymetric floor-area allocation (INSEE FILOSOFI 2019) with 95% uncertainty intervals.

**Results:** Nearly one third of sites (271/878, 30.9%) were Class 1. Around these sites we identified 3,037 residential buildings within 100 m, corresponding to an estimated 17,116 people (95% uncertainty interval 16,952 to 17,280). Sites in direct LoS were closer on average and accounted for most people located within 0–50 m. Patterns varied with urban density, with greater concentration of at-risk sites in less dense contexts.

**Conclusions:** Combining proximity and visibility pinpoints where residents’ potential exposure is most likely. These findings support enforceable minimum setbacks, distance-triggered pre-implementation acoustic study (notably under 100 m), and post-implementation follow-up, and provide actionable inputs for land-use policy (such as integrating distance/LoS screens into plans, conditioning permits, and guiding mitigation) so territorial development proceeds with fewer conflicts.

## 1 Introduction

Padel is a four-player sport played with solid, stringless rackets, using a ball similar to a tennis ball, on a 20 × 10 m court enclosed by glass walls and metal mesh fencing (Martín-Miguel *et al*., 2023). The sport has sustained growth in many countries (International Padel Federation, 2024). In France, the number of participants rose from 110,000 in 2020 to 500,000 in 2025. The French Tennis Federation (FFT), which has held ministerial delegation for the sport since 2014, plans a 170.5% increase in the number of courts (2,218 in 2024) to reach 6,000 by 2029 in order to meet persistent demand (FFT, 2025b).

However, the siting of padel courts, especially in residential, urban, or peri-urban settings, raises coexistence issues with nearby residents (Vieau, 2025). The sport’s acoustic signature, comprising dry, impulsive sounds from ball-racket and ball-glass impacts; metallic clangs on the wire-mesh fence; and players’ vocalizations, makes it particularly disturbing to nearby residents (CidB, 2025b). Multiple studies, reviewed by Leroy T. and Kaiser X., have documented that the sound power of padel activity generally ranges from 89 to 91 dB(A), with peaks up to 102 dB(A) (Leroy, Kaiser and SGS BELGIUM S.A., 2023). This combination produces perceived annoyance higher than for other racket sports such as tennis (Clarke, MacArthur and Whiffin, 2023) and is the leading cause of legal disputes concerning this sport (Gimalac, 2025).

Environmental noise has deleterious effects on the physical and mental health of exposed populations (Basner *et al*., 2014; WHO, 2018; Mehrotra *et al*., 2024; Clark, Vienneau and Aasvang, 2025; Hahad *et al*., 2025). Regular exposure to noise nuisance increases the risk of depression and anxiety by approximately 30% among adults aged 35 years and older (Hu, 2025). Repetitive and impulsive noises, in particular, have psychoacoustic characteristics that make them especially difficult to tolerate (Romito and Fink, 2025) and harmful to health (Yamamura *et al*., 1980; Rajala and Hongisto, 2019; Radun *et al*., 2022). Prolonged exposure to the repetitive, impulsive noises specific to padel has been documented in cases of anxiety-depressive disorders that resulted in criminal convictions in Spain (Audiencia Provincial de Sevilla, Sección 1, 2023). In France, the Hauts-de-France Regional Health Agency (Agence régionale de santé des Hauts-de-France, ARS Hauts-de-France) has added padel activity to the list of developments requiring particular vigilance, alongside shooting ranges, car-wash facilities, and venues broadcasting amplified sound, because of the substantial noise nuisance they can generate (ARS, 2025; Binisti, 2025a).

To prevent noise nuisance from padel courts, some of the countries where the activity is most developed have adopted regulations or recommendations based on setback distances from dwellings. In the Netherlands, a collaborative effort between the Royal Dutch Lawn Tennis and Padel Association (Koninklijke Nederlandse Lawn Tennis Bond, KNLTB), the National Olympic Committee*National Sports Federation (NOC*NSF), the sports-court builders’ association, the association of municipalities, and the Dutch Noise Abatement Foundation produced the”Handreiking Padel en Geluid” (*Padel and Noise Guide*) in January 2023, which now serves as a national reference standard. It recommends a minimum distance of 100 m between padel courts and dwellings, indicates that 160 m is preferable in a quiet residential area, and extends that distance to 210 m when the site comprises four courts (Wadman *et al*., 2021). This recommendation is among the most explicit and quantified internationally and serves as a reference for local authorities and Dutch courts. In Wallonia (Belgium), the Public Service of Wallonia’s guide, based on other acoustic studies, likewise concludes that a distance”beyond 100 m” is necessary (Leroy, Kaiser and SGS BELGIUM S.A., 2023). In France, the FFT’s technical study concludes that beyond 100 m from a dwelling, the risk of nuisance is low; between 75–100 m, the risk is probable; between 50– 75 m, the risk is high and corrective measures are needed; and at <50 m, siting a padel court is”not recommended without substantial acoustic treatment” (Binisti, 2025b; FFT, 2025c). The French Noise Information Center (Centre d’Information sur le Bruit, CidB) reports that the regional health agencies (ARS) recommend a prior acoustic study and, in all cases, avoiding siting padel courts within 50 m of dwellings (CidB, 2025b).

However, in France, municipalities that grant the permits required to install padel courts within their Local Urban Plan (*Plan local d’urbanisme*, PLU) framework (Légifrance, 2001), are not obliged to follow the FFT’s recommendations (Binisti, 2025b) or those issued by the ARS (Binisti, 2025a). As a result, the sport’s expansion, without binding regulatory safeguards, has led to courts being sited close to residential areas, generating environmental nuisances for nearby residents: noise, but also visual and light pollution (Vieau, 2025). Noise nuisance from padel sites located near dwellings has become a national issue, raised in a specific question at the French National Assembly (Spillebout, 2025) and noise control is a stated objective of the National Health-Environment Plan 4 (PNSE 4),”One Environment, One Health” (Gouvernement français, 2021; CidB, 2025a).

To date, no national study in France, or elsewhere, has systematically inventoried padel sites in close proximity to dwellings or quantified the potential scale of the associated noise nuisance. We address this gap with a national spatial analysis of padel sites in metropolitan France to (i) quantify the number that fall below recommended setback thresholds (FFT and ARS) and (ii) estimate the scale and distribution of potential noise-exposure configurations.

Permitting is decided at the municipal level, while regional health agencies issue non-binding opinions and federation documents provide technical guidance without enforceable siting criteria. This heterogeneity motivates a simple, auditable screening framework. In this study, we do not model acoustic propagation nor assess health outcomes. Instead, we provide a national, policy-relevant picture of siting configurations that are plausibly associated with substantial noise nuisance for nearby residents. By quantifying both proximity and line-of-sight (LoS) around padel sites, our results are directly translatable to planning decisions. These findings are intended to inform siting practices rather than to replace technical acoustic assessments. Using proximity as a policy-relevant proxy for potential exposure is consistent with existing regulations and recommendations previously mentioned and planning literatures that rely on distance-based indicators when source-specific measurements are unavailable or impractical at scale (Chakraborty, Maantay and Brender, 2011; Knopper and Ollson, 2011). We extend this logic by pairing simple distance bands with a line-of-sight screen to approximate the presence of intervening built barriers.

## 2 Materials and Methods

### 2.1 Identification and categorization of sites

We compiled a database of active padel sites by aggregating and deduplicating the public”Padel Speak” directory (Padel Speak, 2025) and open government datasets for sports facilities labeled”padel” (Ministère chargé des Sports, 2025). Each site, hosting one or more courts, was geocoded from its address using Google Maps. We used a mixed approach combining quantitative spatial analysis and qualitative review of publicly available information sources to categorize padel sites.

#### Criteria for potential noise exposure

For each padel site, the potential noise exposure of nearby dwellings was classified into three categories using the criteria described below. This classification relied on GIS tools (Google Earth Pro, Google Maps, OpenStreetMap, Géoportail) for distance measurements and topographic review, and was complemented by open imagery (Google Street View; photographs on club websites and social media; press photos).

Each padel site was classified into one of the following three mutually exclusive categories according to these criteria:

#### Class 1: Potentially high exposure

This class groups sites with a high-risk configuration for noise nuisance. A site was assigned here if it met at least one of the two criteria below:

A. Open or semi-covered courts. The distance from the edge of the nearest court to the closest dwelling façade is ≤ 100 m, with no major acoustic screen (continuous, massive building; noise barrier; or significant topographic relief) on the direct line-of-sight (LoS) between the source (court) and the receptor (dwelling).
B. Covered but non-insulated courts. The distance from the structure to the closest dwelling façade is ≤ 50 m, and the structure is lightweight with no evident acoustic insulation (e.g., single-sheet metal panels, bubble-type enclosure, non-masonry shed).

#### Class 2: Potentially low or no exposure

This category includes padel sites where the risk of noise nuisance is considered negligible. A site was assigned to this class if it met at least one of the following criteria:

A. Sufficient setback. The distance from the court (or its enclosing structure) to the closest dwelling façade is > 100 m.
B. Structural acoustic insulation. The court is fully enclosed within a masonry building (e.g., concrete, cinder blocks) or shows clear acoustic treatment (e.g., acoustic sandwich panels), even when the distance is < 100 m.
C. Presence of another, closer dominant noise source than the padel site (e.g., highway, railway).

#### Class 3: Indeterminate exposure

This category is reserved for cases where the available data did not allow a reliable assignment to Class 1 or 2. The reasons for Class 3 were documented and included, in particular:

A. Measurement impossible. Satellite imagery was obscured or outdated, or photographic evidence was unavailable, preventing confirmation of the presence or absence of dwellings within 100 m.
B. Ambiguous acoustic screening. Partial obstacles (e.g., low-rise buildings, discontinuous walls) made it impossible to determine with confidence their sound-attenuation capacity.
C. Uncharacterizable structure. Street View/photos were missing or unusable, preventing determination of the materials and construction of a covered structure.

#### Noise-exposure assessment workflow

Two examiners classified the padel sites in three phases:

1. Phase 1. Independent classification. Each examiner independently reviewed and classified all sites in the database, recording their rationale and supporting evidence in a worksheet.
2. Phase 2. Blind cross-review and re-assessment. Sites with discrepant classifications were flagged. The examiners exchanged worksheets to see each other’s initial label, notes, and evidence. Based on this cross-review and additional independent checks, each examiner either revised or confirmed their own classification, still without consultation.
3. Phase 3. Consensus meeting. Remaining disagreements were isolated and resolved by discussion to assign a consensus class.

Inter-examiner agreement was assessed with Cohen’s kappa (κ) computed from the classification contingency matrix, with 95% confidence intervals (CIs) obtained by bootstrap. To limit observer bias, every site assigned to Class 1 was verified via an automated geometric test in QGIS: the presence of at least one residential building at ≤ 100 m in direct LoS, operationalized as the minimum edge-to-edge distance segment not intersecting any intervening building polygon (line-of-sight/acoustic screen).

### 2.2 Spatial distribution of sites

We examined the regional and municipal distribution of Class 1 padel sites using a standardized over-representation ratio (SIR), defined as Observed/Expected, where Expected = (total number of sites in the region) × (national proportion of Class 1). 95% CIs for SIR were obtained by exact Poisson inversion (Garwood method). Overall heterogeneity was tested with a Pearson χ^2^ test.

Sites were grouped according to INSEE’s communal density grid (INSEE, 2025b), after merging the two rarest categories into a single”Rural (dispersed + very dispersed)” class. Given the ordinal nature of the INSEE categories, we applied a Cochran-Armitage trend test (scores 1→6), weighted by stratum size. 95% CIs for category-specific SIRs were computed using the exact Garwood method. Class 1 proportions by category are reported descriptively.

### 2.3 Exploration of residents’ exposure

The analysis of potential noise exposure for dwellings was carried out in QGIS (v. 3.34 LTS) and focused on Class 1 padel sites. The 2D ground footprint of the reference court (visually selected as the court closest to surrounding dwellings) was modeled by a circular buffer with a 5 m radius centered on the court’s midpoint. This circle is mathematically inscribed within the court’s true footprint (20 × 10 m rectangle), regardless of orientation, making the approach replicable and azimuth-invariant. Because the circle lies inside the real rectangle, the building-to-footprint distance measured to the circle overestimates the distance to the true court edge by up to 5 m along the longitudinal axis (and by 0 m along the transverse axis). All vector data were projected to RGF93 / Lambert-93 (EPSG:2154) to ensure metric accuracy. Data on buildings within 100 m were extracted from IGN’s BD TOPO® (June 2025 edition), which provides, among other attributes, 3D building footprints and a classification of primary use (Institut national de l’information géographique et forestière (IGN), 2025).

To improve the reliability of building-use classification, we conducted a quality-control check to address limitations of BD TOPO. Buildings labeled Unspecified (“Indifférencié”, BD TOPO), potentially dwellings not identified as such, were manually verified using iconographic sources (IGN orthophotography, Google Earth Pro). When confirmed, these buildings were reclassified as Residential (“Résidentiel”). Buildings labeled Residential were retained for the exposure analysis. Non-residential buildings were nevertheless kept in the dataset to evaluate screening effects in the line-of-sight visibility analysis.

#### 2.3.1 Visibility analysis

For each padel site, we performed a 2D proximity analysis in QGIS. First, buildings within 100 m were selected based on the shortest Euclidean edge-to-edge distance between the reference-court footprint and each building footprint (represented as a straight-line segment). We then conducted a line-of-sight analysis to classify buildings: a building was labeled”direct” if the segment was uninterrupted; it was labeled”masked” if any other building, regardless of use or height, intersected that segment (i.e., acted as an intervening screen).

#### 2.3.2 Floor-area-weighted sociodemographic allocation and uncertainty intervals

To characterize exposed populations, for each residential building *b* within INSEE tile *c* (FILOSOFI 2019, carroyage 200 m (INSEE, 2023)), we compute a floor area as *S*_*b*_ = *A*_*b*_ × max(1, *E*_*b*_) (building footprint × total number of floors, including the ground floor; default floor count = 1 if missing). The tile’s total floor area is 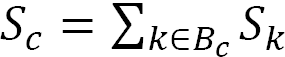, and the allocation weight for building *b* is *P*_*b*_ = *S*_*b*_/*S*_*c*_. INSEE tile totals (individuals, households) are then redistributed in proportion to this weight so that the sum over buildings within a tile exactly matches the observed tile totals.

To quantify allocation uncertainty for a subset *Z* (e.g., *direct* 0–50 m), we compute, for each tile, the share 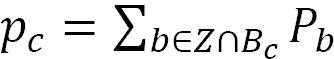, bounded in [0,1]. Assuming individuals (or households) within the tile are randomly assigned according to this share, the exposed count follows *Y*_*c*_ ∼ Binomial(*X*_*c*_, *p*_*c*_) (with *X*_*c*_ = INSEE total for the tile). The overall estimate is 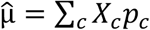 and the variance 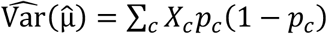 the 95% uncertainty interval (UI) is 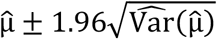 These UIs reflect intra-tile allocation uncertainty, not sampling error.

Distances were summarized at the building level (unweighted) using the mean and the sample standard deviation. Distance bands were 0–50 m and 50–100 m.

Residential buildings located in tiles without available INSEE aggregates (statistical confidentiality) could not be allocated and were excluded from the sociodemographic analysis.

This procedure yielded an enriched dataset in which each exposed building is characterized by its distance to the reference court, its line-of-sight status (*direct* or *masked*), and floor-area-weighted estimates of resident population and households.

##### Sensitivity analyses

We conducted two pre-specified checks: (A) exclusion of buildings within 45–55 m of the court proxy to stress-test the 50 m cut-off, and (B) exclusion of borderline line-of-sight (“grazing”) cases identified by intersecting a +1 m buffer of third-party buildings without intersecting their true polygon after line trimming. All analyses reused the ≤ 100 m inclusion, dasymetric floor-area allocation, and binomial intra-tile 95% uncertainty intervals.

#### 2.3.3 Rationale for policy-oriented metrics

We selected simple, auditable indicators (edge-to-edge distance bands (0–50 m; 50–100 m) and a binary 2D LoS) to enhance policy translatability. Distance bands align with common siting cautions used by local authorities and practitioners, while LoS approximates the presence of intervening built screens. Although these indicators are not substitutes for acoustic modeling, they are reproducible in GIS and can be used as preliminary screening criteria to prioritize projects for further assessment. Our choice of proximity and 2D line-of-sight follows established screening practice. In environmental justice research, distance-to-source metrics are routinely used as reproducible indicators of potential exposure where monitoring is sparse or heterogeneous. Similarly, separation distances and setbacks are widely discussed for other noise-generating land uses (e.g., onshore wind (Knopper and Ollson, 2011)), precisely to provide tractable, policy-translatable criteria short of site-specific modelling. We adopt the same screening logic here and complement it with a basic visibility test to reflect intervening built screens (Chakraborty, Maantay and Brender, 2011).

## 3 Results

### 3.1 Identification and categorization of sites

As of 1 September 2025, our database contained 878 padel sites, each comprising one or more courts. The variables recorded were: club name, address, municipality, department, and region.

Sites were distributed across 747 of metropolitan France’s 34,805 communes, 89 of its 96 departments, and all 13 regions.

The three-phase assessment by the two examiners was conducted over 15 days, from 2 to 16 September 2025. The GIS layers and open imagery consulted correspond to the versions available during this period.

Final distribution (Phase 3, consensus):

- Class 1 (potentially high exposure): 271 sites (30.87%)
- Class 2 (potentially low or no exposure): 567 sites (64.58%)
- Class 3 (indeterminate exposure): 40 sites (4.56%)

Inter-examiner agreement at Phase 1 was 91.23% (κ = 0.83; 95% CI [0.79–0.87]), and all remaining discrepancies were resolved in the subsequent phases. For all Class 1 sites, QGIS measurements confirmed the presence of at least one residential building within 100 m with direct LoS (i.e., no intervening built screens; see end of”Spatial distribution of sites” section below).

### 3.2 Spatial distribution of sites

Table 1 shows that the regional distribution of Class 1 padel sites is not homogeneous in France (χ^2^ = 24.85; p = 0.0156). Provence-Alpes-Côte d’Azur accounts for 14.24% of all sites nationally but 22.88% of Class 1 sites; the within-region proportion classified as Class 1 is 49.6% (SIR = 1.61; 95% CI [1.35–1.87]). Conversely, Grand Est (SIR = 0.58; 95% CI [0.30–0.89]) and Centre-Val de Loire (SIR = 0.22; 95% CI [0.00– 0.67]) are under-represented..

**Table 1.** Distribution of padel sites by region in metropolitan France.

| Region | Padel sites (n) | National share of sites (%) | Class 1 sites (n) | Within-region Class 1 (%) | National share of Class 1 sites (%) | Class 1 SIR (over-representation ratio) (95% CI) | Regional pop. density (inh./km²) |
| --- | --- | --- | --- | --- | --- | --- | --- |
| Provence-Alpes-Côte d’Azur | 125 | 14.24% | 62 | 49.60% | 22.88% | 1.61 [1.35–1.87] | 166.9 |
| Auvergne-Rhône-Alpes | 128 | 14.58% | 43 | 33.59% | 15.87% | 1.09 [0.84–1.34] | 118.5 |
| Occitanie | 153 | 17.43% | 50 | 32.68% | 18.45% | 1.06 [0.85–1.27] | 85.3 |
| Normandie | 29 | 3.30% | 9 | 31.03% | 3.32% | 1.01 [0.46–1.55] | 111.7 |
| Pays de la Loire | 39 | 4.44% | 12 | 30.77% | 4.43% | 1.00 [0.56–1.48] | 122.7 |
| Nouvelle-Aquitaine | 123 | 14.01% | 34 | 27.64% | 12.55% | 0.90 [0.65–1.13] | 73.7 |
| Île-de-France | 81 | 9.23% | 22 | 27.16% | 8.12% | 0.88 [0.58–1.19] | 1036.5 |
| Bourgogne-Franche-Comté | 20 | 2.28% | 5 | 25.00% | 1.85% | 0.81 [0.18–1.45] | 58.5 |
| Bretagne | 37 | 4.21% | 9 | 24.32% | 3.32% | 0.79 [0.36–1.26] | 127.8 |
| Hauts-de-France | 55 | 6.26% | 11 | 20.00% | 4.06% | 0.65 [0.33–1.00] | 187.8 |
| Grand Est | 67 | 7.63% | 12 | 17.91% | 4.43% | 0.58 [0.30–0.89] | 96.5 |
| Corse | 6 | 0.68% | 1 | 16.67% | 0.37% | 0.54 [0.00–1.65] | 41.5 |
| Centre-Val de Loire | 15 | 1.71% | 1 | 6.67% | 0.37% | 0.22 [0.00–0.67] | 65.9 |
| All regions (Metropol. France) | 878 | 100% | 271 | 30.87% | 100% |  |  |

*Population density (inhab./km<sup>2</sup>) : INSEE source, « Estimations de population au 1<sup>er</sup> janvier 2025 » (INSEE, 2025a). Values computed/reported using the current regional boundaries.*

**Table 2.** Distribution of padel sites by INSEE’s communal population density grid.

| INSEE communal density category | Sites (n) | National Share of sites (%) | Class 1 sites (n) | Within-category Class 1 (%) | National share of Class 1 sites (%) | Class 1 SIR (over-representation ratio) (95% CI) |
| --- | --- | --- | --- | --- | --- | --- |
| Large urban centers | 276 | 31.44% | 72 | 26.09% | 26.57% | 0.85 [0.70–0.99] |
| Intermediate urban centers | 181 | 20.62% | 59 | 32.60% | 21.77% | 1.06 [0.86–1.25] |
| Urban belts | 153 | 17.43% | 43 | 28.10% | 15.87% | 0.91 [0.70–1.13] |
| Rural towns | 139 | 15.83% | 43 | 30.94% | 15.87% | 1.00 [0.77–1.24] |
| Small towns | 79 | 9.00% | 34 | 43.04% | 12.55% | 1.39 [1.06–1.74] |
| Rural (dispersed + very dispersed) | 50 | 5.69% | 20 | 40.00% | 7.38% | 1.30 [0.88–1.73] |
| All INSEE categories (Metropol. France) | 878 | 100% | 271 | 30.87% | 100% |  |

*The INSEE categories “Rural dispersed” and “Rural very dispersed” were merged.*

The spatial distribution of sites across France is shown in Figure 1. Site locations were aggregated to the centroid of their respective communes. Point displacement symbology was used to visualize multiple sites in the same commune.

**Figure 1.**
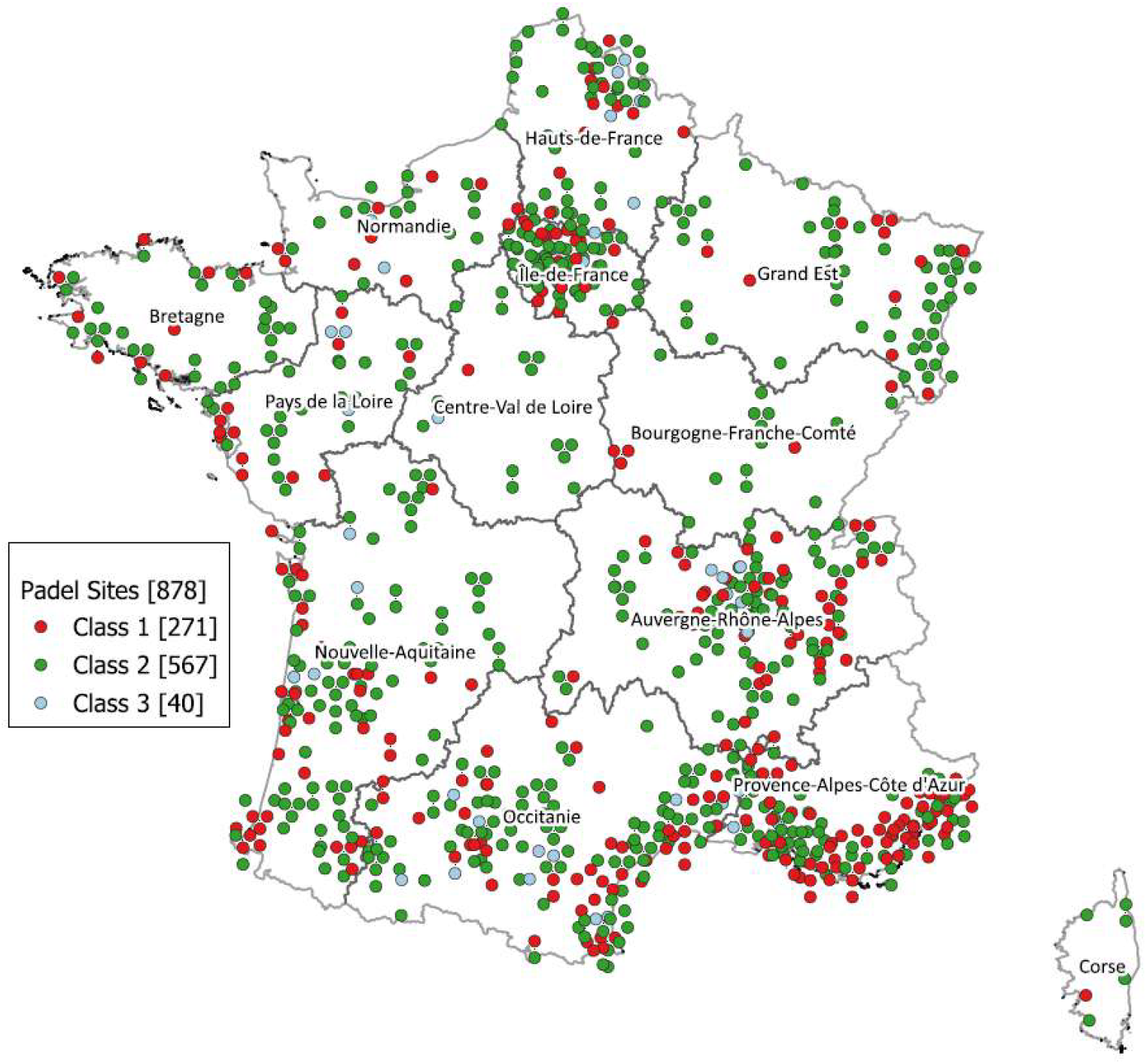
Geographic distribution of padel sites across metropolitan France’s regions.

On INSEE’s 6-level communal density grid (INSEE, 2025b), the distribution of Class 1 padel sites shows overall heterogeneity (χ^2^ = 11.20; p = 0.0476). A significant trend also emerges along the density gradient: the proportion of Class 1 sites increases as density decreases (Cochran-Armitage trend test: Z = 2.66; p = 0.0078). The”Small towns” category is significantly over-represented (SIR = 1.39; 95% CI [1.06–1.74]).

For the 271 Class 1 sites, we counted 534 outdoor courts, 37 covered but non-enclosed courts, and 21 courts housed within lightweight enclosed structures. Projecting a 5 m circular buffer around the midpoint of the court closest to dwellings, then computing edge-to-edge distances to buildings labeled Residential in BD TOPO, revealed inconsistencies for 11 sites: the minimum distance to a residential building exceeded 100 m, which did not match the situation observed by the examiners. Review indicated these discrepancies stemmed from incomplete labeling in BD TOPO. To correct this bias, 32 buildings initially classified as Unspecified (“Indifférencié”, BD TOPO) were reclassified as Residential (“Résidentiel”) after manual verification using GIS sources (Google Earth Pro satellite imagery and IGN aerial orthophotography). These reclassified buildings were then included in the final building set.

### 3.3 Exploration of residents’ exposure

The dataset comprises 7,471 buildings within 100 m of 271 padel sites. Figure 2 shows an example of a 2D QGIS display of buildings within 100 m of a padel court (circular buffer). Each building is described by its distance to the site (straight-line segment), its visibility status (solid = direct; dashed = masked), and the sociodemographic indicators of its INSEE FILOSOFI 2019 tile (not shown in Figure 2). Of these buildings, 3,097 are residential (red, orange, sand, yellow) and 4,374 non-residential (gray), the latter retained solely to characterize visibility/masking. Residential buildings are split into 1,379 with direct LoS and 1,718 masked.

**Figure 2.**
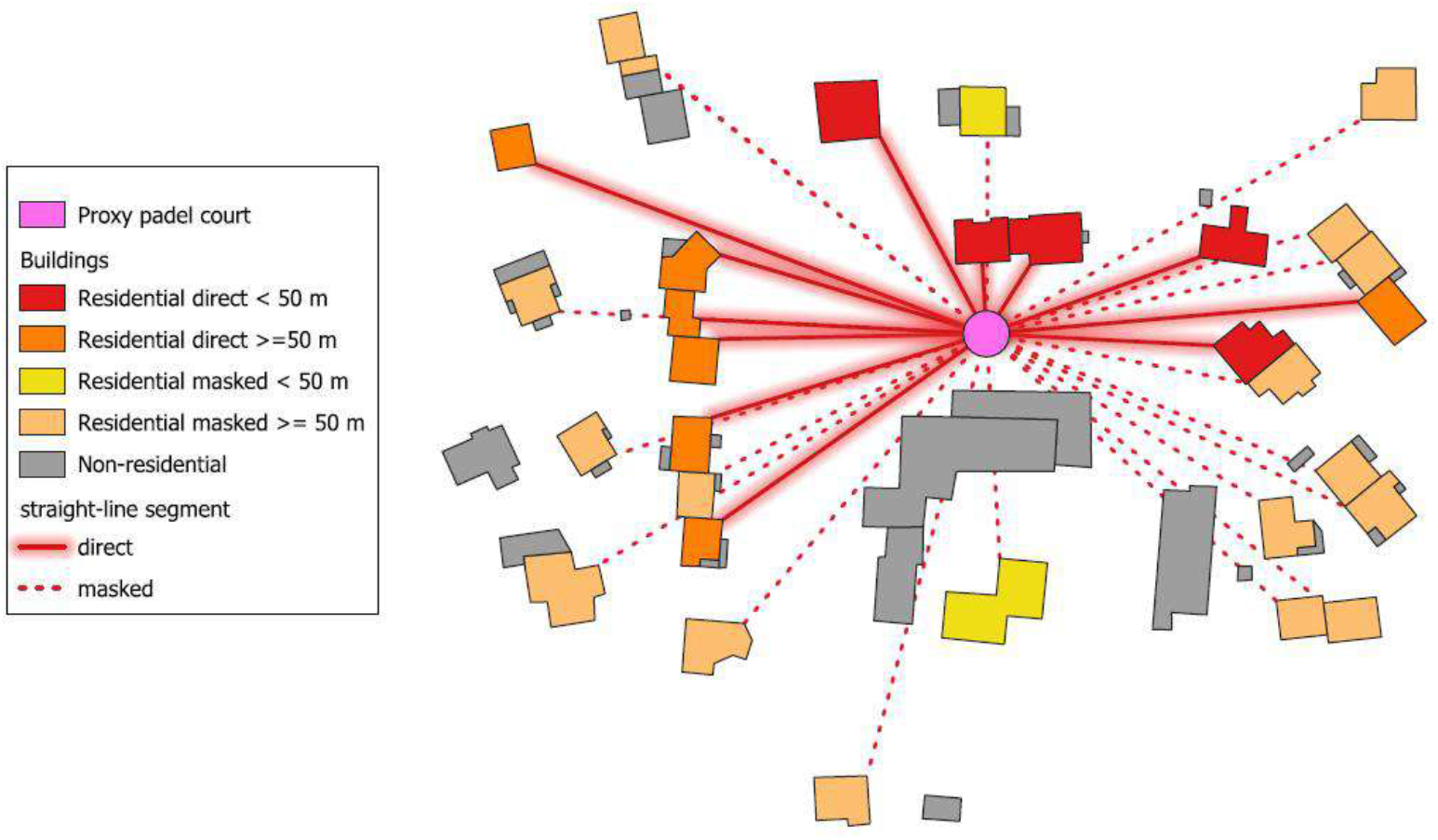
Example of a 2D QGIS representation of buildings within 100 m of a padel court.

**Figure 3.**
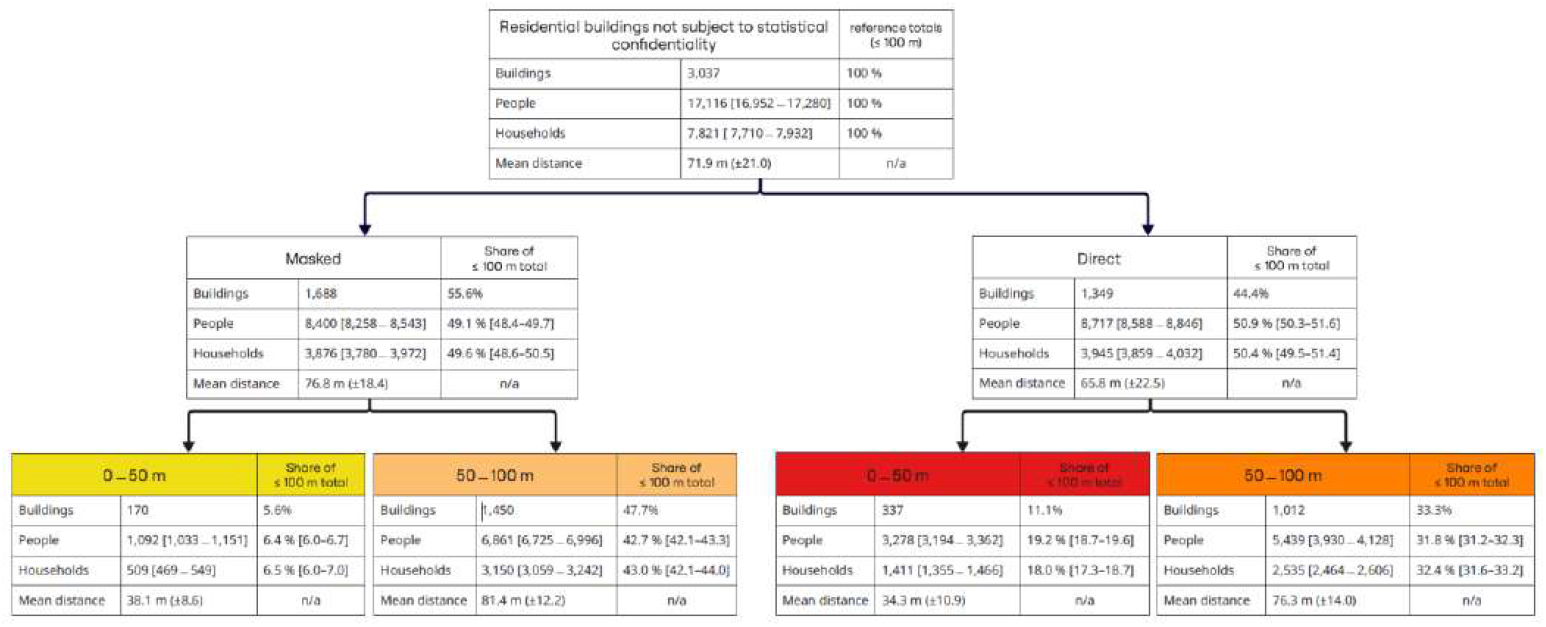
Hierarchical summary of residential buildings within 100 m of Class 1 padel sites, by visibility (direct vs masked) and distance band (0–50 m; 50–100 m).

However, the sociodemographic analyses (Figure 2) are based on a final dataset of 3,037 residential buildings with complete information and 608 INSEE tiles; tiles subject to statistical confidentiality were excluded from allocation, resulting in the exclusion of 60 residential buildings (≈ 1.9% of the initial 3,097). No cases of invalid tile floor area (≤ 0) were observed.

Among the 3,037 residential buildings not subject to statistical confidentiality, 44.4% have direct LoS and 55.6% are masked. Overall, the floor-area allocation yields an estimated 17,116 people (95% UI [16,952–17,280]) and 7,821 households (95% UI [7,710–7,932]) within 100 m of the installation’s reference court, with a mean distance of 71.9 m (±21.0).

Direct buildings account for 8,717 people (95% UI [8,588–8,846]) and 3,945 households (95% UI [3,859– 4,032]), at a shorter mean distance (65.8 m, ±22.5) than masked buildings (76.8 m, ±18.4). The share of buildings located at 0–50 m is roughly doubled under direct visibility (11.1%) vs masked (5.6%), and the majority of people at 0–50 m are in direct buildings (3,278/4,370 ≈ 75%).

#### Sensitivity analyses

Option B (excluding”grazing” LoS) reduced totals modestly (buildings 2,834; people 16,480; households 7,527), preserving the visibility-distance pattern. Option A (excluding 45–55 m) yielded 2,783 buildings, 15,264 people, and 6,962 households. The combined Option A+B gave 2,595 buildings, 14,678 people, and 6,692 households (95% UIs in Table S3, supplemental materials).

## 4 Discussion

The close siting of some padel sites near dwellings, combined with an acoustic signature of impulsive, repetitive sounds, exposes nearby residents to noise nuisance that may adversely affect quality of life. Our national analysis shows that the distance-visibility coupling structures potential exposure: direct line-of-sight configurations are more often associated with very short distances and concentrate the most sensitive situations. In a context where recommendations exist but are unevenly known or implemented, these findings argue for clearer siting conditions. Our approach aligns with proximity-based methods long used to flag potential environmental nuisances and inequities, where distance functions as a conservative screen pending detailed assessment. Evidence from other planning domains shows that setback frameworks can guide siting and mitigation decisions even when direct health effects are not evaluated (Chakraborty, Maantay and Brender, 2011; Knopper and Ollson, 2011).

To our knowledge, while GIS-population approaches exist for other noise sources (e.g., road traffic) (Kephalopoulos *et al*., 2014), they have never been applied to padel. To date, there is no peer-reviewed national study that characterizes resident exposure around padel sites using a spatial approach coupled with sociodemographic allocation. Available bibliometric overviews place padel research largely in the fields of play analysis, performance, and biomechanics, with environmental nuisance and built-environment proximity not treated as primary objects of study (Denche-Zamorano *et al*., 2024). Our work therefore fills a gap by documenting, at the national scale, potential exposure configurations and the order of magnitude of the populations concerned.

At the scale of metropolitan France, our work shows that about one third of padel sites do not meet recommended setback distances, suggesting noise-nuisance exposure for thousands of nearby residents. Moreover, the over-representation of the most problematic sites in certain density categories (notably”Small towns”) and the shorter distances observed under direct line-of-sight converge on the idea that the distance-visibility coupling is a major determinant of potential exposure. These findings arise in a regulatory and guidance landscape (France, the Netherlands, Belgium) that is heterogeneous and, in practice, does not ensure effective control of siting near housing.

Our observations are consistent with existing recommendations and technical guides that advocate minimum distances (often ≥100 m, modulated by the number of courts and the urban context) precisely to limit the impact of the activity’s impulsive, repetitive sounds. The fact that some sites lie below these benchmarks helps explain the frequently reported mismatch between the sport’s rapid growth and the rise in neighborhood complaints. In this context, our spatial approach provides a national quantification of a previously undocumented phenomenon and pinpoints the highest-risk configurations, useful for prioritizing actions (prevention, mediation, urban planning and noise nuisance mitigation) (Conseil National du Bruit, 2011; Gouvernement français, 2021).

The overall heterogeneity and the significant trend along the density gradient (from dense urban to rural) suggest that urban-integration constraints (built fabric, available land, and building screens) and the degree of land-use leeway modulate the likelihood of”too-close” sitings. In dense urban environments, most courts are indoors, housed in solid, acoustically insulated buildings, which is why we did not classify them as Class 1. In lower-density settings, building screens are less common and outdoor courts are more frequent, exposing dwellings at short distances; this helps explain SIR > 1 in certain categories. As expected, direct LoS is accompanied by shorter distances and concentrates the majority of people very close (0–50 m). Viewed through the psychoacoustics of impulsive sounds, this distance-visibility coupling strengthens the plausibility of greater disturbance, independent of the sheer number of dwellings affected.

### Forces

Our database lists 878 active padel sites, designed to be exhaustive as of 1 September 2025, whereas the FFT reported 834 in metropolitan France over the same period (FFT, 2025a). The study benefits from national coverage, a documented, reproducible GIS workflow, and a cautious two-examiner classification, including an Indeterminate class when information was ambiguous or missing, plus formal spatial verification via QGIS distance and line-of-sight calculations, which limits the risk of false positives. The reported 95% uncertainty intervals (UIs) for counts (people, households) stem from intra-tile statistical allocation; they are imputation margins, not sampling error. Sensitivity analyses indicate that our conclusions are robust to borderline visibility classification (Option B), and that the 50 m split is not driving the results spuriously, although excluding the 45–55 m band (Option A) unsurprisingly lowers counts near the threshold. Operational implications therefore hinge primarily on the distance-visibility configuration rather than edge-case geometry.

### Limitations

We did not conduct acoustic measurements or propagation modeling, and we did not evaluate health outcomes. Our indicators (distance bands and 2D LoS) are therefore screening proxies intended to inform siting and prioritization for further assessment. Our database coverage, though broad, may still be incomplete, leading to under-ascertainment of sites. The approximation of the proxy court footprint (5 m inscribed circle) may overestimate building distances and thus undercount buildings in the 0–50 m and 50–100 m bands; any bias is limited to cases lying within 5 m of the band thresholds. The visibility analysis relies on a 2D model (shortest Euclidean distance between entities) without 3D modeling (altimetry, heights, materials), which may over- or underestimate actual screening. For multi-court sites, using a single proxy court may understate cumulative exposure. Sociodemographic data (INSEE FILOSOFI 2019) are projected onto the 2025 building stock; local changes (construction/demolition, population movements) may introduce temporal mismatch. We used dasymetric allocation proportional to floor area; alternative schemes (footprint-only, different occupancy assumptions, inclusion of structural attributes) could have been considered and may locally alter redistributed totals. The reported 95% UIs reflect intra-tile allocation uncertainty. Application of statistical confidentiality led to the exclusion of some buildings, causing localized underestimation despite prorated allocation on other tiles. Finally, we did not model contextual factors (traffic, ambient noise, usage rules), and our exposure indicators (distance-visibility) remain indirect, warranting caution for any health inference.

Operationally, the results support a clearer framework for siting conditions: minimum setback distances modulated by configuration (number of courts, built context), enhanced review for projects < 100 m from dwellings, and tailored mitigation (effective barriers, enclosure/acoustic treatment, hours-of-use management). Mapping direct line-of-sight situations at very short distances provides a pragmatic lever to prioritize interventions.

Finally, several research avenues emerge: coupling 3D geometry and acoustic-propagation modeling with multi-period in-situ measurements; linking these exposures to indicators of annoyance and health (housing turnover/sales, complaints, sleep disturbance, mental health) using existing sources or ad-hoc data collected in experimental settings (ISO, 2021); and evaluating the effectiveness and cost-benefit of mitigation measures to inform operational decisions.

## 5 Conclusion

Our results document a gap between recommendations and certain padel site implementations near housing. Our national maps of proximity and LoS highlight where current siting practices are most likely to generate noise-related conflicts, providing a practical basis for distance-triggered acoustic assessment, context-modulated setbacks, and targeted mitigation. In a context of rapid growth, this represents both a public-health concern and a spatial-planning challenge. Three priorities emerge:

1. an enforceable national framework (distance/typology thresholds, modulated by number of courts and urban context);
2. a pre-implementation acoustic study using a reproducible protocol for every new project, incorporating in situ measurements and usage scenarios;
3. post-implementation follow-up (complaints, measurements, mediation) to adjust problematic configurations through appropriate engineering solutions (effective barriers, enclosure/acoustic treatment of structures), and, if necessary, relocation of sites.

Taken together, the distance and visibility criteria provide an operational framework to reconcile the sport’s expansion with the protection of nearby residents.

## Supporting information

Table 3 Sensitivity analysis

## 6 Data availability

Aggregated datasets supporting this study are available on Zenodo (concept DOI: 10.5281/zenodo.17290320). Versioned releases will include any subsequent code and documentation updates.

## 7 Competing interests

This research was conducted jointly as part of the corresponding author’s academic activities and the national citizen collective NAMP, a non-profit organization of individuals affected by nuisances associated with padel or pickleball activities. All authors are members of these entities. They declare no other competing interests.

## 8 Ethics

No human participants or animals were involved. We used publicly available spatial data; ethics approval was not required.

## 9 Funding

No specific funding was received.

## 10 Acknowledgments

The authors thank the members of the NAMP collective for their support and Professor Jean Gaudart for helpful discussions and methodological advice.

## 11 Statement on the use of AI tools

During the preparation of this work the authors used ChatGPT 5 Pro (OpenAI) and Gemini Pro (Google) for translating some sentences. After using this tool/service, the authors reviewed and edited the content as needed and take full responsibility for the content of the published article.

## Notes

### Summary of Updates

References update and style reformat Improve impact on policy Improve methodology justification

