## Supplementary material for "Padel Sites in Metropolitan France: Spatial and Sociodemographic Characterization of Residents’ Potential Noise Exposure": Table 3 Sensitivity analysis

**Table S3. Sensitivity analyses (distance–visibility), metropolitan France, ≤100 m**

Four scenarios are reported: Baseline (main analysis), Option A (exclusion of buildings at 45–55 m), Option B (exclusion of borderline line-of-sight “grazing” cases), and Option A+B (A and B combined). Socio-demographic estimates are obtained by dasymetric floor-area allocation; 95% uncertainty intervals (UI) reflect intra-tile allocation uncertainty (binomial scheme).

**Totals by scenario (≤100 m)**

| Scenario | Buildings (n) | People (95% UI) | Households (95% UI) |
| --- | --- | --- | --- |
| Baseline | 3,037 | 17,117 (95% UI 16,916–17,318) | 7,822 (95% UI 7,686–7,958) |
| Option A | 2,783 | 15,264 (95% UI 15,073–15,455) | 6,962 (95% UI 6,833–7,091) |
| Option B | 2,834 | 16,480 (95% UI 16,283–16,677) | 7,527 (95% UI 7,395–7,659) |
| Option A+B | 2,595 | 14,678 (95% UI 14,491–14,865) | 6,692 (95% UI 6,566–6,818) |

**Breakdown by cell (visibility × distance band, ≤100 m)**

| Scenario | Visibility | Band | Buildings  (n; %) | People  (n; 95% UI; %) | Households  (n; 95% UI; %) | Mean distance (m; ± SD) |
| --- | --- | --- | --- | --- | --- | --- |
| Baseline | Direct (LoS) | 0–50 | 337; 11.1% | 3,278; 95% UI 3,194–3,362; 19.1% | 1,411; 95% UI 1,355–1,466; 18.0% | 34.3 ± 10.9 |
| Baseline | Direct (LoS) | 50–100 | 1,012; 33.3% | 5,439; 95% UI 5,335–5,543; 31.8% | 2,535; 95% UI 2,464–2,606; 32.4% | 76.3 ± 14.0 |
| Baseline | Masked | 0–50 | 170; 5.6% | 1,092; 95% UI 1,033–1,151; 6.4% | 509; 95% UI 469–549; 6.5% | 38.1 ± 8.6 |
| Baseline | Masked | 50–100 | 1,518; 50.0% | 7,308; 95% UI 7,170–7,446; 42.7% | 3,367; 95% UI 3,274–3,460; 43.0% | 81.1 ± 13.4 |
| Option A | Direct (LoS) | 0–50 | 273; 9.8% | 2,485; 95% UI 2,410–2,559; 16.3% | 1,046; 95% UI 996–1,095; 15.0% | 31.1 ± 9.7 |
| Option A | Direct (LoS) | 50–100 | 931; 33.5% | 5,167; 95% UI 5,066–5,268; 33.9% | 2,419; 95% UI 2,350–2,487; 34.7% | 78.4 ± 12.6 |
| Option A | Masked | 0–50 | 129; 4.6% | 751; 95% UI 703–800; 4.9% | 347; 95% UI 314–381; 5.0% | 35.1 ± 7.6 |
| Option A | Masked | 50–100 | 1,450; 52.1% | 6,861; 95% UI 6,725–6,996; 44.9% | 3,150; 95% UI 3,059–3,242; 45.2% | 82.4 ± 12.2 |
| Option B | Direct (LoS) | 0–50 | 325; 11.5% | 3,236; 95% UI 3,153–3,319; 19.6% | 1,392; 95% UI 1,337–1,447; 18.5% | 34.2 ± 10.9 |
| Option B | Direct (LoS) | 50–100 | 923; 32.6% | 5,112; 95% UI 5,012–5,212; 31.0% | 2,381; 95% UI 2,313–2,448; 31.6% | 75.8 ± 14.0 |
| Option B | Masked | 0–50 | 153; 5.4% | 1,050; 95% UI 992–1,108; 6.4% | 489; 95% UI 450–529; 6.5% | 38.2 ± 8.4 |
| Option B | Masked | 50–100 | 1,433; 50.6% | 7,082; 95% UI 6,946–7,218; 43.0% | 3,265; 95% UI 3,173–3,356; 43.4% | 81.1 ± 13.4 |
| Option A+B | Direct (LoS) | 0–50 | 262; 10.1% | 2,447; 95% UI 2,373–2,521; 16.7% | 1,029; 95% UI 980–1,078; 15.4% | 31.0 ± 9.7 |
| Option A+B | Direct (LoS) | 50–100 | 845; 32.6% | 4,865; 95% UI 4,769–4,962; 33.1% | 2,278; 95% UI 2,213–2,344; 34.0% | 78.0 ± 12.6 |
| Option A+B | Masked | 0–50 | 116; 4.5% | 719; 95% UI 671–767; 4.9% | 332; 95% UI 300–365; 5.0% | 35.3 ± 7.5 |
| Option A+B | Masked | 50–100 | 1,372; 52.9% | 6,647; 95% UI 6,513–6,780; 45.3% | 3,053; 95% UI 2,963–3,143; 45.6% | 82.4 ± 12.2 |

**Notes.** “Direct (LoS)”: unobstructed line of sight; “Masked”: line of sight interrupted by a third-party building. “Grazing” cases are distance lines that touch a +1 m buffer of another building without intersecting its true polygon (after end trimming). Percentages are computed within the ≤100 m perimeter.
